# Effect of Nirmatrelvir/Ritonavir (Paxlovid) on Hospitalization among Adults with COVID-19: an EHR-based Target Trial Emulation from N3C

**DOI:** 10.1101/2023.05.03.23289084

**Authors:** Abhishek Bhatia, Alexander J. Preiss, Xuya Xiao, M. Daniel Brannock, G. Caleb Alexander, Robert F. Chew, Megan Fitzgerald, Elaine Hill, Elizabeth P. Kelly, Hemalkumar B. Mehta, Charisse Madlock-Brown, Kenneth J. Wilkins, Christopher G. Chute, Melissa Haendel, Richard Moffitt, Emily R. Pfaff, The N3C Consortium

## Abstract

This study leverages electronic health record data in the National COVID Cohort Collaborative’s (N3C) repository to investigate disparities in Paxlovid treatment and to emulate a target trial assessing its effectiveness in reducing COVID-19 hospitalization rates. From an eligible population of 632,822 COVID-19 patients seen at 33 clinical sites across the United States between December 23, 2021 and December 31, 2022, patients were matched across observed treatment groups, yielding an analytical sample of 410,642 patients. We estimate a 65% reduced odds of hospitalization among Paxlovid-treated patients within a 28-day follow-up period, and this effect did not vary by patient vaccination status. Notably, we observe disparities in Paxlovid treatment, with lower rates among Black and Hispanic or Latino patients, and within socially vulnerable communities. Ours is the largest study of Paxlovid’s real-world effectiveness to date, and our primary findings are consistent with previous randomized control trials and real-world studies.

## Introduction

The COVID-19 pandemic has had a profound global impact, with over 761 million cases and 6.8 million deaths as of 21 March, 2023. ^1^ This crisis has been met with research and drug development efforts at unprecedented speed, resulting in a number of new treatments aimed at lessening the risk of progression to severe disease. One such treatment, nirmatrelvir/ritonavir (Paxlovid), is a combination protease inhibitor that blocks the replication of SARS-CoV-2. In December 2021, the US Food and Drug Administration (FDA) issued an Emergency Use Authorization (EUA) for Paxlovid, enabling its prescription to high-risk SARS-CoV-2-positive patients aged 12 and older. ^2^ The EUA was based on the phase II-III EPIC-HR trial, which reported an 88.9% reduction in the risk of COVID-19-related hospitalization or death among those who received Paxlovid compared to those who received placebo. ^3^

Since the EUA issuance, a few studies have assessed the effectiveness of Paxlovid using real-world data. An electronic health record (EHR)-based study in the Kaiser Permanente Southern California health system found that fewer than 1% of patients treated with Paxlovid (*n* = 5, 287) were hospitalized or seen in the emergency department within 5-15 days of the drug being dispensed, though this was not compared with a control group. ^4^ Another study leveraging a large repository of Israeli health care data found a 46% reduction in risk of severe COVID-19 outcomes in patients treated with Paxlovid (*n* = 4, 737) when compared with controls, showing a protective effect, but at a lower magnitude than the original EPIC-HR analysis. ^5^ A retrospective cohort study in Hong Kong also found that patients treated with Paxlovid (*n* = 4, 921) were at decreased risk of hospitalization, with a weighted hazard ratio of 0.79. ^6^ Although these studies offer valuable preliminary evidence of Paxlovid’s real-world effectiveness, there remains a dearth of research specifically aimed at understanding the causal effects of the drug on COVID-19 outcomes with large, representative samples derived from real-world data.

A number of EHR studies have also used real-world data to uncover racial and social disparities in the prescription of various COVID-19 treatments in the United States, including Paxlovid. Prior to Paxlovid’s authorization, multiple studies noted racial and social disparities among SARS-CoV-2-positive patients in access to and treatment with monoclonal antibodies (mAb), with Black and Hispanic or Latino patients less likely than White patients to receive treatment with mAb. ^7–9^ A more recent large-scale study of EHR data revealed that those disparities have persisted with Paxlovid; from April through July of 2022, the rate of Paxlovid treatment was 35.8% lower among Black adult patients than White adult patients. ^10^

Through the National Institute of Health’s (NIH) National COVID Cohort Collaborative (N3C), we have the opportunity to replicate and expand on these and other recent analyses of Paxlovid efficacy and treatment patterns. We use the target trial emulation (TTE) framework and a large, geographically and demographically diverse cohort from N3C’s EHR data repository. ^11^ Here, we characterize the population prescribed Paxlovid, assess potential disparities in Paxlovid prescription, and estimate the causal effect of Paxlovid use on hospitalization among SARS-CoV-2-positive adults in the United States. ^11^

## Results

The hypothesized target trial protocol is articulated in Table 1, and we emulate each component to define our base population within the N3C database (See Figure 1).

**Table 1.** Protocol of a Target Trial to Estimate the Effect of Paxlovid on the rate of hospitalization in the 28-days following a positive SARS-CoV-2 test

| Protocol Component | Description under Target Trial Conditions | Method of Target Trial Emulation in this study |
| --- | --- | --- |
| Eligibility criteria | Persons aged 18 and older who have one or more risk factors for severe COVID-19 as per CDC guidelines and who have had an COVID-19 infection during the study period.<sup>31</sup> | Persons aged 18 and older who have one or more risk factors for severe COVID-19 as per CDC guidelines and who have had either a documented COVID-19 diagnosis, or laboratory confirmed positive SARS-CoV-2 test during the study period.<sup>31</sup> |
| Treatment strategies | Complete course of treatment with Paxlovid within five days of symptom onset. | Complete course of treatment with Paxlovid within five days of the date of laboratory-confirmed SARS-CoV-2 test, or COVID-19 diagnosis. |
| Assignment procedures | Participants will be randomly assigned to treatment or control at baseline and will be aware of their treatment assignment. | Coarsened exact matching on pre-treatment covariates to ensure exchangeability of treatment and control groups and emulate random assignment. |
| Follow-up period | 28 days following a positive SARS-CoV-2 test | 28 days following either positive SARS-CoV-2 test, or COVID-19 diagnosis |
| Outcome | Hospitalization within follow-up period | Hospitalization within follow-up period |
| Causal contrasts | Intention-to-treat effect | Intention-to-treat effect |
| Analysis plan | Intention-to-treat effect estimated via comparison of hospitalization rates among individuals assigned to each treatment arm, adjusting for pre-treatment covariates. Additional analyses to include the main and interaction effects of vaccination. | Coarsened exact matching on pre-treatment covariates; Intention-to-treat effect estimated via comparison of hospitalization rates among individuals in each treatment arm in the matched cohort, adjusting for pre-treatment covariates. Additional analyses to include the main and interaction effects of vaccination. |

**Fig. 1.**
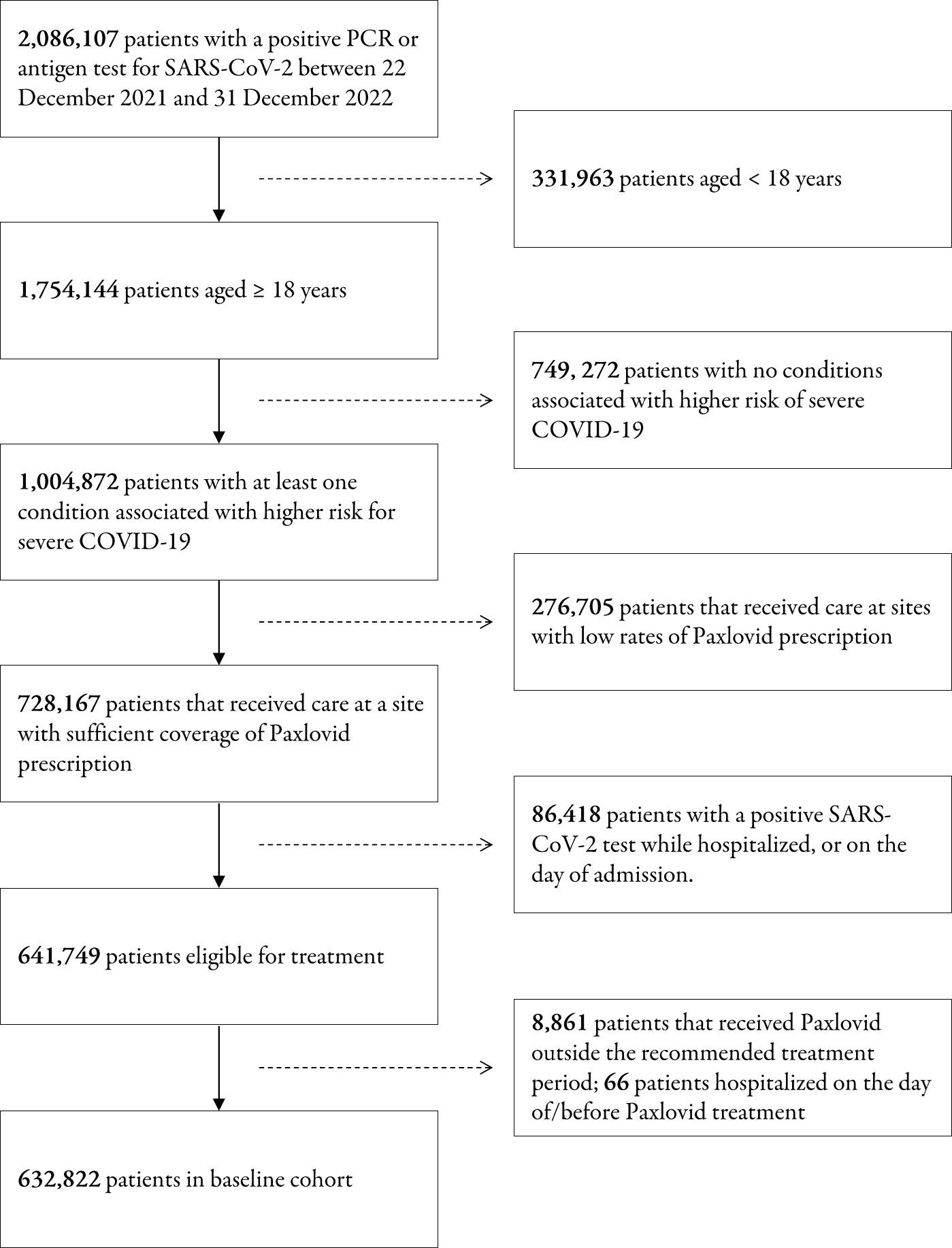
Proportion of individuals in base population stratified by Race/Ethnicity, and ZIP Code-level Community Wellbeing Index (CWBI)

### Patient Characteristics

In the unmatched cohort (within our defined study period), a total of 632,822 patients had a valid COVID-19 index date during the study period, of which 111,443 (17.6%) were treated with Paxlovid, and 698 (0.6%) were hospitalized. After applying the eligibility criteria to the patient population and study sites, a total of 33 of 76 study sites were retained. Patients were matched across treatment groups by characteristics selected *a priori* associated with the treatment assignment and outcome, yielding an effective sample size of 410,642 across 33 sites, balanced across all covariates. The characteristics of all patients during the study period are presented in Table 2, stratified by treatment group.

**Table 2.** Baseline Population Characteristics Before and After Matching

| Characteristic | Before Matching |  | After Matching |  |
| --- | --- | --- | --- | --- |
|  | No Paxlovid<br>(N= 521,379) | Paxlovid<br>(N=111,443) | No Paxlovid<br>(N=306,132) | Paxlovid<br>(N=104,510) |
| Hospitalization<sup>1</sup> |  |  |  |  |
| No | 510,706 (98.0%) | 110,745 (99.4%) | 300,583 (98.2%) | 103,882 (99.4%) |
| Yes | 10,673 (2.0%) | 698 (0.6%) | 5,549 (1.8%) | 628 (0.6%) |
| Sex |  |  |  |  |
| Female | 315,078 (60.4%) | 67,597 (60.7%) | 189,642 (61.9%) | 63,914 (61.2%) |
| Male | 206,198 (39.5%) | 43,829 (39.3%) | 116,490 (38.1%) | 40,596 (38.8%) |
| Missing | 103 (0.0%) | <20 (0.0%) | - | - |
| Age (in years) |  |  |  |  |
| 18-24 | 15,963 (3.1%) | 1,429 (1.3%) | 3,757 (1.2%) | 1,018 (1.0%) |
| 25-34 | 35,194 (6.8%) | 4,538 (4.1%) | 11,921 (3.9%) | 3,657 (3.5%) |
| 35-49 | 70,172 (13.5%) | 12,716 (11.4%) | 34,950 (11.4%) | 11,172 (10.7%) |
| 50-64 | 215,304 (41.3%) | 42,112 (37.8%) | 127,144 (41.5%) | 40,101 (38.4%) |
| 65+ | 184,746 (35.4%) | 50,648 (45.5%) | 128,360 (41.9%) | 49,562 (47.4%) |
| Missing | - | - | - | - |
| Race/Ethnicity<sup>2</sup> |  |  |  |  |
| Asian (NH) | 11,082 (2.1%) | 3,594 (3.2%) | 3,393 (1.1%) | 2,544 (2.4%) |
| Black or African American (NH) | 74,907 (14.4%) | 10,627 (9.5%) | 27,638 (9.0%) | 9,457 (9.0%) |
| Hispanic or Latino (Any Race) | 49,908 (9.6%) | 8,332 (7.5%) | 18,638 (6.1%) | 7,021 (6.7%) |
| White (NH) | 335,385 (64.3%) | 81,891 (73.5%) | 235,601 (77.0%) | 80,081 (76.6%) |
| Other (NH) | 5,716 (1.1%) | 920 (0.8%) | 978 (0.3%) | 502 (0.5%) |
| Unknown | 44,381 (8.5%) | 6,079 (5.5%) | 19,884 (6.5%) | 4,905 (4.7%) |
| Charlson Comorbidity Index |  |  |  |  |
| 0 | 194,689 (37.3%) | 46,596 (41.8%) | 126,679 (41.4%) | 44,822 (42.9%) |
| 1-2 | 188,703 (36.2%) | 42,488 (38.1%) | 119,336 (39.0%) | 40,479 (38.7%) |
| 3-4 | 54,101 (10.4%) | 11,228 (10.1%) | 29,086 (9.5%) | 9,914 (9.5%) |
| 5-10 | 32,788 (6.3%) | 5,513 (4.9%) | 15,113 (4.9%) | 4,648 (4.4%) |
| 11+ | 3,644 (0.7%) | 523 (0.5%) | 638 (0.2%) | 296 (0.3%) |
| Missing | 47,454 (9.1%) | 5,095 (4.6%) | 15,280 (5.0%) | 4,351 (4.2%) |
| Community Wellbeing Index<sup>3</sup> |  |  |  |  |
| 0-45 | 2,646 (0.5%) | 275 (0.2%) | 327 (0.1%) | 157 (0.2%) |
| 46-55 | 152,670 (29.3%) | 26,978 (24.2%) | 91,926 (30.0%) | 25,470 (24.4%) |
| 56-65 | 193,631 (37.1%) | 46,639 (41.9%) | 106,456 (34.8%) | 43,891 (42.0%) |
| 66-100 | 26,644 (5.1%) | 9,961 (8.9%) | 13,750 (4.5%) | 8,879 (8.5%) |
| Missing | 145,788 (28.0%) | 27,590 (24.8%) | 93,673 (30.6%) | 26,113 (25.0%) |
| Month of COVID-19 diagnosis |  |  |  |  |
| December 2021 | 39,670 (7.6%) | 77 (0.1%) | - | - |
| January 2022 | 166,565 (31.9%) | 1,544 (1.4%) | 902 (0.3%) | 64 (0.1%) |
| February 2022 | 37,600 (7.2%) | 931 (0.8%) | 69,739 (22.8%) | 1,495 (1.4%) |
| March 2022 | 11,775 (2.3%) | 845 (0.8%) | 16,468 (5.4%) | 865 (0.8%) |
| April 2022 | 16,902 (3.2%) | 3,805 (3.4%) | 4,226 (1.4%) | 725 (0.7%) |
| May 2022 | 38,567 (7.4%) | 12,984 (11.7%) | 10,782 (3.5%) | 3,420 (3.3%) |
| June 2022 | 39,959 (7.7%) | 14,950 (13.4%) | 31,292 (10.2%) | 12,227 (11.7%) |
| July 2022 | 45,957 (8.8%) | 19,455 (17.5%) | 32,867 (10.7%) | 14,135 (13.5%) |
| August 2022 | 39,646 (7.6%) | 16,128 (14.5%) | 39,158 (12.8%) | 18,580 (17.8%) |
| September 2022 | 26,454 (5.1%) | 11,101 (10.0%) | 32,965 (10.8%) | 15,303 (14.6%) |
| October 2022 | 20,185 (3.9%) | 8,386 (7.5%) | 20,819 (6.8%) | 10,338 (9.9%) |
| November 2022 | 18,149 (3.5%) | 9,131 (8.2%) | 15,889 (5.2%) | 7,766 (7.4%) |
| December 2022 | 19,950 (3.8%) | 12,106 (10.9%) | 14,412 (4.7%) | 8,275 (7.9%) |

<sup>1</sup>Any hospital admission in the 28-days following a positive SARS-CoV-2 test result<sup>2</sup>abbrev. NH = Non-Hispanic.<sup>3</sup>Community Wellbeing Index (CWBI) is a measure of five inter-related community-level domains, with higher CWBI values corresponding to a higher level of protective community-level social determinants of health. 32

Among the unmatched cohort, there were large, statistically significant differences in assignment to Paxlovid treatment - 12.42% of Black, non-Hispanic patients and 14.31% of Hispanic patients were treated with Paxlovid, compared to 19.63% of White, non-Hispanic patients, and 24.49% of Asian, non-Hispanic patients (*χ*^2^ test of independence p-value *<* 0.001). When stratified by patients’ residential areas, patients who lived in areas with higher Community Wellbeing Index (CWBI) values (and lower corresponding social vulnerability) were also more likely to be treated with Paxlovid. (*χ*^2^ test of independence p-value *<* 0.001). (See Figure 2)

**Fig. 2.**
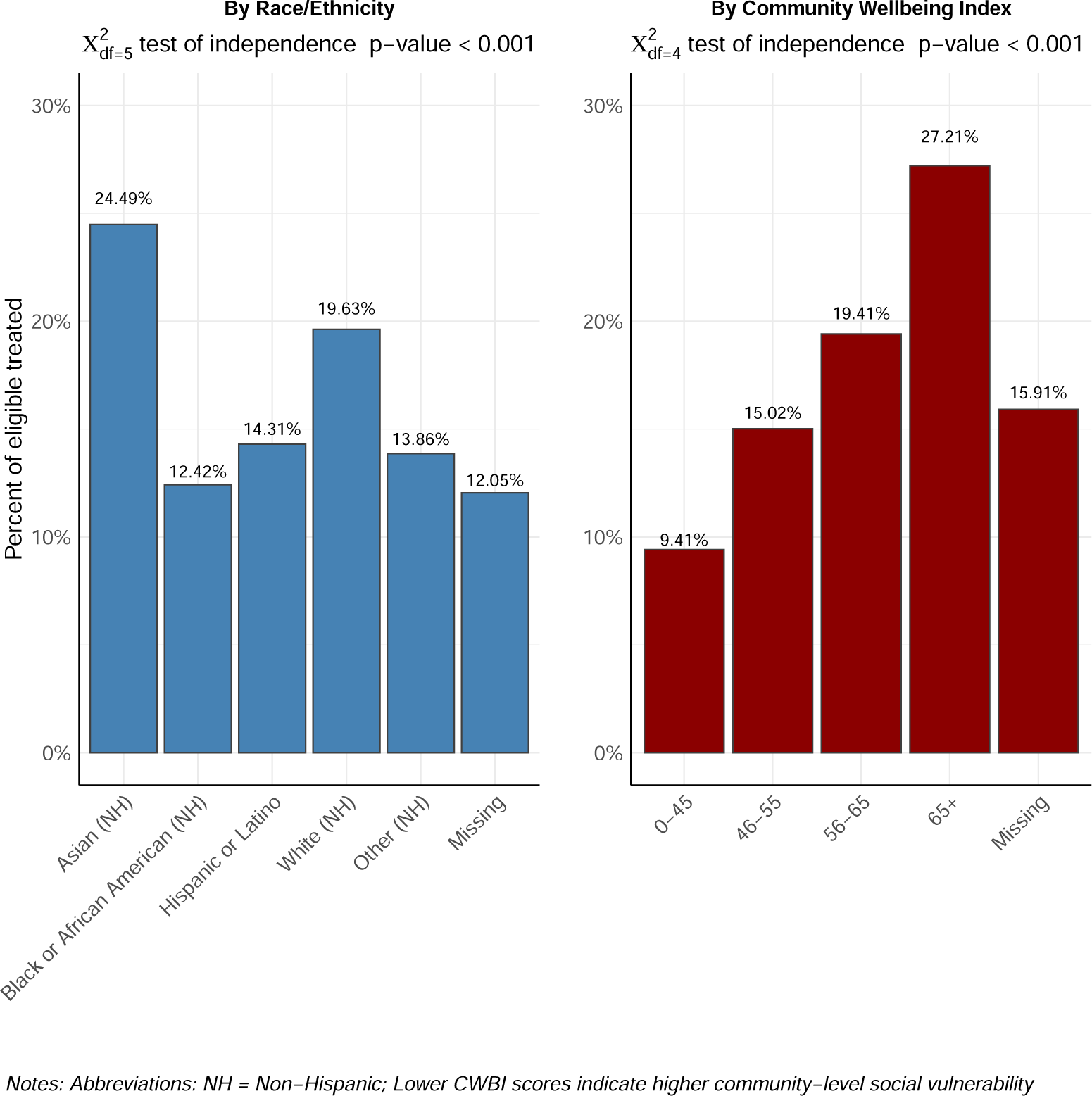
Proportion of individuals in base population stratified by Race/Ethnicity, and ZIP Code-level Community Wellbeing Index (CWBI)

### Effect of Paxlovid on Hospitalization

In the unmatched cohort, 698 (0.6%) patients treated with Paxlovid were hospitalized in the follow-up period, compared to 10,673 (2.0%) patients in the untreated group. In the primary analysis using the matched cohort, patients treated with Paxlovid had lower unadjusted causal odds of hospitalization compared with patients not treated with Paxlovid (OR, 0.33; 95% CI, 0.24-0.45), and this was consistent with the estimate adjusted for potential confounders (OR, 0.35; 95% CI, 0.29-0.42). (See Table 3)

**Table 3.** Odds of hospitalization in Paxlovid-treated vs. Non-Paxlovid-treated patients

|  | *Dependent variable: Hospitalization* |  |  |
| --- | --- | --- | --- |
|  | Unadjusted (1) | Adjusted (2) | Vaccine-adjusted (3) |
|  | OR (95% CI) | OR (95% CI) | OR (95% CI) |
| Paxlovid Treatment |  |  |  |
| No (control) | *ref* | *ref* | *ref* |
| Yes (treatment) | 0.33 (0.24-0.45)*** | 0.35 (0.29-0.42)*** | 0.32 (0.24-0.42)*** |
| Vaccination Status<sup>1</sup> |  |  |  |
| Unvaccinated | - | - | *ref* |
| Vaccinated | - | - | 0.49 (0.41-0.58)*** |
| Vaccination*Paxlovid<sup>2</sup> | - | - | 1.08 (0.71-1.64) |
| *Observations* | 410,642 | 410,642 | 136,815 |
| *Log Likelihood* | -31590.1 | -25466.1 | -6551.1 |
| *Akaike Inf. Crit.* | 63184.0 | 51000.2 | 13172.3 |

<sup>1</sup>Unvaccinated refers to patients who received 0 doses at index, Vaccinated refers to patients who recieved atleast 2 doses at least 14 days prior to index.
<sup>2</sup>Interaction term
Note: Model (2) adjusts for sex, age, race and ethnicity, Charlson Comorbidity Index, Community-well being index, data partners, and month of COVID-19 Index date. Model (3) additionally adjusts for the main and interaction of Vaccination.
$p < .05^*$ , $p < .01^{**}$ , $p < .001^{***}$

The adjusted estimates in the primary analysis adjust for the effects of CWBI captured at the patient level, including patients that were categorized as “Missing” for any CWBI data, and we assume that missingness was unrelated to unmeasured confounding or treatment assignment. Results of our sensitivity analysis showed that the estimates for our primary analysis were robust against missing CWBI data, both when sites were dropped (OR, 0.36; 95% CI, 0.29-0.44), or when the covariate itself was dropped (OR, 0.37; 95% CI, 0.31-0.45). Additionally, sensitivity analysis aimed at identifying the potential effects of immortal time bias also showed that our estimates were stable (OR, 0.38; 95% CI, 0.31-0.46). (Supplementary Table 1).

### Effect adjusted by COVID-19 Vaccination Status

For our subgroup analysis estimating the effect of Paxlovid on hospitalization adjusting for the effect of vaccination, individuals from nine sites with trusted vaccine data were included. A total of 170,063 individuals included in the base population were vaccinated, of which 45,929 (27.0%) received Paxlovid, and 1,873 (1.1%) were hospitalized; 51,306 individuals were unvaccinated, of which 6,633 (12.9%) received Paxlovid, and 1,152 (2.2%) were hospitalized. After CEM matching (*n* = 136, 815), weighting, and adjusting for both the main effects and interaction effects of vaccination status, this analysis was consistent with the primary results, and patients prescribed Paxlovid had lower adjusted causal odds of hospitalization (OR, 0.32; 95% CI, 0.24-0.42). Vaccinated patients also had lower adjusted causal odds of hospitalization (OR, 0.49; 95% CI, 0.41-0.58). Additionally, there was no significant evidence of an interaction effect between the effect of Paxlovid and vaccination on hospitalization at 28 days, suggesting no heterogeneity by vaccination status in the effect of Paxlovid treatment (OR, 1.08; 95% CI, 0.73-1.61).

## Discussion

Despite the widespread use of nirmatrelvir/ritonavir (Paxlovid) for the treatment of COVID-19, we are still in the early stages of understanding its protective effect at scale in real-world settings. In this target trial emulation using the N3C database, one of the largest longitudinal observational datasets on COVID-19 patients in the United States, individuals treated with Paxlovid within five days of a COVID-19 diagnosis or positive SARS-CoV-2 test had 65% lower odds of hospitalization, compared with those never treated with Paxlovid. When accounting for vaccination status, individuals treated with Paxlovid had a 68% lower odds of hospitalization, and this effect did not depend on an individual’s vaccination status. We also find significant differences in distribution of Paxlovid across race, ethnicity, and social vulnerability strata. Paxlovid is thus effective at lessening the risk of hospitalization, but is not equitably distributed.

We examined the distribution of Paxlovid treatment by race and ethnicity, along with a measure corresponding to a patient’s residential ZIP Code-level community well being index (CWBI). We found large differences in Paxlovid treatment rates by race/ethnicity and CWBI. Across race/ethnicity strata, Black and Hispanic/Latino patients were on average less likely to receive Paxlovid treatment, compared to White and Asian patients. Additionally, patients from communities with higher levels of social vulnerability (lower CWBI values) were also less likely to receive Paxlovid treatment. Our findings are consistent with prior research from December 2021-July 2022 that also documented disparities in outpatient oral antiviral treatment by both ZIP Code-level social vulnerability and race/ethnicity, and showed that Black patients had 35.8% lower rates of treatment compared to White patients, and Hispanic patients had 29.9% lower rates of treatment compared to non-Hispanic patients. These disparities in access to treatment are of particular concern due to the growing evidence base that shows that individuals in these groups are more likely to experience higher levels of COVID-19 exposure, discrimination in access to care, and severe COVID-19 health outcomes. ^12–17^ The reasons for these disparities are a complex constellation of factors including (and not limited to) a history of systemic discrimination and racism in treatment, lack of physical and economic resources to facilitate equal access in vulnerable communities, lack of information and knowledge about treatment options, and language barriers owing to a lack of culturally competent care. ^10^ Without attention, recognition, and remediation on the part of providers, public health agencies, the health system, and communities, the disproportional burden of COVID-19 will only further exacerbate existing health inequities in the United States.

Findings from our primary analysis were consistent with existing randomized control trial results. The EPIC-HR randomized control trial (*n* = 2, 246) among symptomatic, non-hospitalized, high-risk patients assessed the effect of Paxlovid on hospitalization or death through a 28-day follow-up period, and found a 89.9% risk reduction. ^4^ The lower magnitude of effect in our findings may be attributable to differences in study design (as noted by Najjar-Debbiny et. al): 1) differences in dominant strains at the time of the study; 2) their inclusion of only symptomatic patients versus our inclusion of all COVID-19 positive patients (inclusive of asymptomatic patients); 3) their earlier treatment assignment within five days from symptom onset versus our treatment assignment within five days of a COVID-19 diagnosis or a laboratory-confirmed SARS-CoV-2 test; and 4) our intention-to-treat approach with no information on adherence to treatment, compared to their clinical trial setting. ^5^ We also adjusted for the effects of vaccination, and interestingly, we found that the effects of treatment with Paxlovid were not heterogeneous across vaccinated and unvaccinated individuals. This finding is of particular interest given the preliminary evidence from the EPIC-SR trial that the impact of Paxlovid on vaccinated populations was uncertain. ^18^ Further research is needed on the effectiveness and cost-effectiveness of Paxlovid treatment in these populations. It is also worth noting that the omission of vaccination status did not significantly bias our estimates in our primary analysis.

Our findings are also consistent with other real-world analyses suggesting a protective effect of Paxlovid. Najjar-Debbiny et. al applied Cox hazard models and found that treatment with Paxlovid resulted in a 46% reduced risk of severe COVID-19 and 80% reduced risk of mortality in Israel, with no evidence for the interaction effect of Paxlovid and vaccination (*n* = 180, 351). ^5^ While their outcome of interest was severe COVID-19 or death, ours examined hospitalization, complementing their findings and notably, expanding on their regression-adjusted approach by including a causal inference framework through target trial emulation to estimate the causal effect of Paxlovid. Our estimates also support results from a retrospective cohort study in Hong Kong (*n* = 93, 833), which estimated a propensity score-weighted 21% reduction in hospitalization risk among patients treated with Paxlovid. ^6^ However, while this analysis used population-level vaccination coverage as an estimate for patient-level vaccination status, our sub-analysis matched patients on their health-record documented vaccination information to provide a more precise estimate among vaccinated populations.

### Strengths

This study has several strengths that underscore the value of large-scale EHR repositories for advancing comparative effectiveness research. In the absence of large-scale RCTs, target trial emulations allow researchers to explore treatment effects in real-world settings and identify the most effective treatments for a variety of health conditions. This study is one of a few studies that apply methods to emulate hypothetical target trials, accounting for the effect of confounding. ^19–22^ The estimated effects were also consistent across sensitivity analyses targeted at addressing issues in both missingness and potential immortal time bias. Additionally, the analyses were conducted using a large, comprehensive database of EHR data from 33 contributing sites across the United States, increasing generalizability, and decreasing the potential for issues that typically arise from misclassification in administrative or claims data. ^23^ The volume of data contained in the N3C database allowed for coarsened exact matching while preserving statistical power, instead of weighting approaches that may be sensitive to model misspecification and the potential for extreme weights leading to biased estimates. ^24;25^

### Limitations

This study also has a few limitations. First, since the data did not include any further information on the dose received over the study period (Paxlovid is typically given orally twice daily for five days), we assume complete adherence to treatment and no loss-to-follow-up, such that the intention-to-treat effect is equal to the per-protocol effect. Second, the sub-analysis on vaccinations did not include individuals with incomplete courses of vaccination (1 dose), nor information on the timing of vaccination relative to COVID-19 infection, and therefore we were unable to shed light on whether the response to Paxlovid varied by additional strata of vaccination. Third, the outcome of interest was a discrete measure of hospitalized/not hospitalized, and further studies may benefit from taking a time-to-hospitalization approach and modeling the effectiveness of Paxlovid using a Cox proportional hazards model, which may further inform smaller differences in treatment-response. Due to the many possible patterns over time of COVID-19 index, Paxlovid treatment, and hospitalization, determining time zero (*t*_0_) in this study was not trivial. We opted to treat hospitalization as a discrete outcome in our follow-up period to simplify the study’s concept of time. Related to the difficulty in establishing *t*_0_, while our sensitivity analysis aimed to demonstrate that our estimates would be consistent even in the presence of substantial immortal time bias, further sensitivity analyses using a nested target trial emulation framework may be useful in substantiating this. ^26;27^ Fourth, it is well-documented that EHRs are susceptible to missing data when patients do not seek care, care is provided outside of the reporting facility, or a condition is documented outside of the structured EHR (e.g., in clinical notes), and it is likely that our estimates may be biased if missingness was related to any residual unobserved confounding. ^28–30^ We took several steps to mitigate the risk of missing data – all individuals in our cohort have established care at the partner facility both before and after their acute COVID-19 event, as evidenced by documented healthcare encounters, and our vaccination sub-analysis is limited to facilities with a high recorded vaccine ratio to reduce the number of individuals misclassified as unvaccinated. Fifth, our inclusion criteria of Paxlovid treatment within five days of COVID-19 index differs from the indication of treatment within five days of symptom onset. However, we note that within our base cohort, 95.70% of treated patients were treated within one day of COVID-19 index. Finally, our study is subject to the assumptions of all causal inference studies: consistency, positivity, and exchangeability. In particular, the assumption of exchangeability rests on the assumption that there are no unmeasured confounders.

## Conclusion

Among patients with COVID-19 in our study period, the odds of hospitalization within a 28-day follow-up period was 65% lower in patients treated with Paxlovid within five days of COVID-19 index, compared with patients who were never treated with Paxlovid. Although there remains the potential for unmeasured confounding, the results of our large-scale study using EHR data are consistent with the evidence base of smaller-scale RCTs and smaller real-world data studies across other geographies. Our results demonstrate the potential for further research using the target trial emulation framework with observational data to supplement clinical trials. We also found disparities in the rates of Paxlovid treatment. Black patients, Hispanic or Latino patients, and patients living in more vulnerable communities were treated with Paxlovid at a significantly lower rate than others. Taking action to remediate these disparities will equalize the opportunity for all high-risk patients to prevent severe COVID-19 outcomes.

## Data Availability

All data is available in the N3C Data Enclave to those with an approved protocol and data use request from an institutional review board. Data access is governed under the authority of the National Institutes of Health; more information on accessing the data can be found at https://covid.cd2h.org/for-researchers. See Haendel et. al. for additional detail on how data is ingested, managed, and protected within the N3C Data Enclave.

## Methods

We performed a target trial emulation to assess the effect of Paxlovid treatment within five days of COVID-19 index on the risk of hospitalization within 28 days of COVID-19 index (see Eligibility Criteria). We followed a two-step process for emulating target trials with observational data: first, we articulated the causal question of interest in the form of a hypothetical randomized trial protocol, specifying eligibility criteria, treatment strategies, treatment assignment, the study period for follow-up, the outcome of interest, causal contrasts, and the analysis plan to estimate effects. ^33^ Second, we emulated each component of this protocol using patient-level data inside the NIH-hosted N3C Secure Data Enclave, which integrates EHR data for 18 million patients across 76 participating sites across the United States. N3C’s methods for patient data acquisition, ingestion, and harmonization have been reported in detail elsewhere. ^34–36^ All analyses and results as part of this study are reported in adherence with the Strengthening the Reporting of Observational Studies in Epidemiology (STROBE) reporting guidelines. ^37^

### Eligibility Criteria

We defined our study period as December 23, 2021, the day after FDA authorization of Paxlovid, to December 31, 2022. To meet the eligibility criteria for the study as per the target trial protocol, we specified the following inclusion criteria: 1) having a documented COVID-19 index date within the study period (with index date defined as the earliest date of either (a) a COVID-19 diagnosis or (b) a positive SARS-CoV-2 test result), 2) being *≥* 18 years of age as of the COVID-19 index date, 3) having one or more risk factors for severe COVID-19 as per CDC guidelines, including age *≥* 50 years old, or the presence of underlying medical conditions associated with a conclusive higher risk of severe COVID-19. ^31^ We excluded all patients *<* 18 due to the potential for differences in both clinical characteristics and prescription practices among pediatric and adult patients prescribed Paxlovid. ^38;39^

Additionally, since we were interested in quantifying the causal effect of Paxlovid treatment on the outcome of hospitalization, we applied three exclusion criteria to exclude: 1) patients who were hospitalized on or before the COVID-19 index date or date of treatment with Paxlovid (outcome precluding treatment), 2) patients who received Paxlovid before their COVID-19 index date, and 3) patients who received Paxlovid after the recommended five days following their COVID-19 index date. In order to ensure that data were captured from sites with high fidelity and adequate coverage, we only included data from sites with at least 1% of eligible patients, and a minimum of 100 patients, treated with Paxlovid during the study period.

### Defining Treatment and Outcome

Eligible patients were categorized by their treatment exposure, defined as having been treated with Paxlovid within five days of their COVID-19 index date, with controls defined as patients never treated with Paxlovid. We selected an eligibility window of five days in adherence with recommended clinical guidelines, and to minimize heterogeneity and potential for indication bias. For patients who were never treated with Paxlovid, we used the date of their earliest indication of COVID-19 (diagnosis or positive lab result) within the study period as the index date. For those in the treatment group, we used their earliest indication of COVID-19 within five days of their first Paxlovid treatment date in the study period. As per our exclusion criteria, we excluded patients that received Paxlovid after the 5-day eligibility period. Within the N3C enclave, the “Paxlovid or nirmatrelvir” concept set was used to identify drug exposures that correspond to Paxlovid (10 Observational Medical Outcomes Partnership [OMOP] concepts). ^40^ We followed patients for a 28-day period following their COVID-19 index date. Our primary outcome of interest was hospitalization at any point in time during the 28-day follow-up period, specified as a discrete measure corresponding to any hospitalization (*Y* = 1) or no hospitalization (*Y* = 0). For the purposes of this analysis, we did not consider time-to-hospitalization, severity at the time of admission, or any downstream outcomes such as in-hospital death or discharge.

## Statistical Analysis

### Overview

First, we applied a two-sided Chi-squared test to examine the distribution of Paxlovid treatment across two covariates: 1) patient race/ethnicity, and 2) a ZIP Code-level Community Wellbeing Index (hereafter referred to as CWBI). The CWBI measure is a composite index of social determinants of health available within the N3C database, with higher CWBI values corresponding to a higher level of protective community-level social determinants of health. The index methodology was developed by Sharecare and the Boston University School of Public Health. CWBI values are derived from the patient’s residential ZIP Code-level data across five key inter-related community-level domains: healthcare access (ratios of healthcare providers to population), resource access (libraries and religious institutions, employment, and grocery stores), food access (access to grocery stores and produce), housing and transportation (home values, ratio of home value to income, and public transit use), and economic security (rates of employment, labor force participation, health insurance coverage rate, and household income above the poverty level). ^32^

Next, we used a potential outcomes framework to compare the rate of hospitalization among patients who received Paxlovid during the five days following a positive SARS-CoV-2 test to those who did not. To adjust for confounding and emulate random assignment in the context of the target trial, we matched individuals in the treatment and control cohorts on a set of pre-treatment variables: sex, age (binned), race and ethnicity, Charlson Comorbidity Index (CCI) value (as a proxy for all underlying medical conditions; binned), CWBI (binned), month of COVID-19 onset, and site of care provision, including all data present in their electronic health record as of their positive SARS-CoV-2 test. Due to the prevalence of literature suggesting disparity in treatment assignment and outcomes by race, ethnicity, and other social determinants of health, we include these measures as potential confounders. ^7–10^ Sex, age, and comorbidities are known to affect both care seeking for and the outcome of COVID-19. The index month was included because Paxlovid treatment rates, viral variants, and infection rates changed during the study period. CCI was coded as missing when no condition exposures were present in N3C prior to index. CWBI was coded as missing when patient ZIP Code was not reported.

We matched patients in the treatment and control groups using coarsened exact matching (CEM) and weighted observations by their CEM weights. CEM weights were defined as:

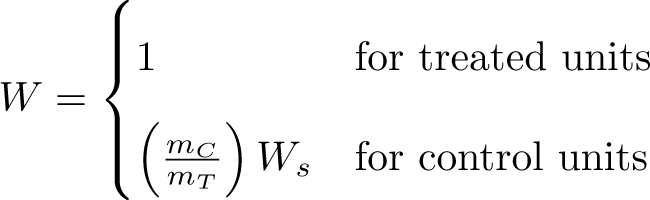

*W_s_* for control units where *m_C_* and *m_T_* are the numbers of control units and treated units in the sample, 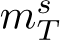 and 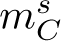 are the number of control units and treated units in stratum *s*, and 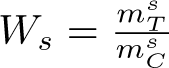.

CEM offers several advantages over other matching methods. ^41^ Furthermore, when the likelihood of treatment is heavily determined by the covariates (as in this study), weighting methods like inverse probability of treatment weighting tend to assign extreme weights to treated patients for whom treatment was very unlikely (and vice versa). CEM avoids this issue. Bins used to coarsen covariates were based on subject matter knowledge of the measurement scale of each variable (e.g., age groups based on strata with known differences in COVID-19 outcomes). We examined the results of the specified coarsened exact matching to ensure that it yielded a balanced cohort with a sufficiently large effective sample size.

### Estimation

Our primary analysis aims to estimate the total effect of receiving treatment with Paxlovid within five days of COVID-19 index date (treatment), compared to not receiving Paxlovid (control), on the discrete outcome of hospitalization.

The unadjusted average treatment effect on the treated (ATT) was first estimated using a difference-in-means estimator:

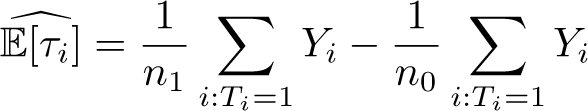

The unadjusted causal odds ratio among the matched population was then estimated as:

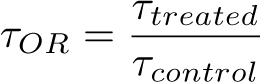

The adjusted treatment effect was calculated using a CEM-weighted mixed-effects logistic regression model with a discrete response variable corresponding to hospitalization (*Y* = 1, *Y* = 0), controlled for independent pre-treatment variables. We include site-specific random intercepts to account for within-cluster homogeneity in outcomes and to estimate the influence of the cluster on the outcomes of the individuals within the cluster. We assume that the fixed effects are invariant across clusters and therefore do not include random slopes in the model.

The final CEM-weighted mixed-effects logistic regression model is specified as:

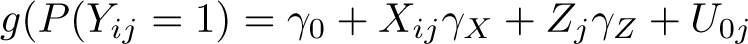

where *γ*_0_ is the intercept, *γ_X_* are the effects of the covariates at the individual level, *γ_Z_* are the effects of the observed covariates at the cluster level, and *g* denotes the logit link function. The cluster-level random intercepts *U*_0*j*_ are assumed to have and are uncorrelated with the included covariates; these induce a data-partner-specific interpretation desired for odds ratios. ^42^

### Vaccination-adjusted Subanalysis

Using the same method applied to the primary analysis described above, we also conducted a subanalysis with COVID-19 vaccination status included as a covariate. We considered this important for two reasons. First, we hypothesized that vaccination status may be a confounder of Paxlovid treatment and hospitalization, largely through the latent variable of infection severity. If so, our primary analysis would violate the assumption of no unmeasured confounding. Second, we hypothesized that Paxlovid treatment may be less effective among vaccinated patients. There is mixed evidence for this dynamic. Pfizer’s EPIC-SR trial found no significant treatment effect among vaccinated patients with one risk factor for severe COVID-19. ^43^ A large health system found that Paxlovid treatment further reduced the likelihood of hospitalization and death among vaccinated patients, but that the treatment effect was smaller among fully vaccinated patients. ^18^

For this subanalysis, we used a modified cohort of patients from sites with reliable information on patient vaccination status. Vaccination status in N3C is subject to misclassification, particularly among patients who were vaccinated outside of data partner systems. We determined the subset of sites with reliable patient-level vaccination records by the ratio of two statistics: (1) the proportion of individuals who are documented as vaccinated in their EHR and (2) the proportion of individuals who are truly vaccinated. We calculated the first statistic from the EHR data for each site. We estimated the second statistic using CDC-reported vaccination rates for the counties served by each partner facility. ^44^ CDC vaccination rates by county are included as a data asset in N3C. Patient counties were inferred using a county-ZIP crosswalk. Each patient’s likelihood of vaccination was drawn from their county’s vaccination rate and the overall expected vaccination rate for a partner facility was computed as the mean of their patients’ vaccination likelihood. We defined a facility’s recorded vaccine ratio as the ratio of these two statistics and limited the vaccination sub-analysis to individuals from facilities with a ratio of at least 0.66. ^45^

Accordingly, we categorized patients by their vaccination status prior to their COVID-19 index date, defined as having completed a full course of vaccination at least 14 days prior to index. Partially vaccinated patients and patients who were fully vaccinated fewer than 14 days prior to index were excluded from the analysis. With treatment, follow-up period, and outcomes consistent with the specification of the primary analysis, we re-estimated the treatment effect of Paxlovid on hospitalization using the previously specified mixed-effects logistic regression model, this time with the additional adjustment for the independent effect of vaccination status, and the interaction effect of vaccination and treatment with Paxlovid.

### Sensitivity Analysis

We conducted two sensitivity analyses with regard to our primary analysis. First, we assessed the sensitivity of our estimated treatment effects to the CWBI as a matching variable, as well as the potential impact of missing CWBI data on treatment effect estimates (if the missingness was related to unmeasured confounders or treatment assignment). CWBI was the only covariate with an overall missing rate (27.40%) higher than 10 percent. CWBI missingness was largely determined by site. Six sites do not report patient ZIP Code data to N3C. An additional four sites had missing ZIP Code data for more than 10 percent of patients. To test for sensitivity to this, we removed data from all sites with a 10% or higher missingness rate for the CWBI covariate, then repeated the matching process and re-estimated the treatment effects on the outcome. In addition, we also repeated the analysis by excluding the CWBI variable entirely from both the matching and estimation processes and reported the resulting adjusted effect estimates. Second, we aimed to understand the potential immortal time bias that may be present in our primary analysis. In our primary analysis, we define time zero (*t*_0_) as the date of a patient’s COVID-19 index date. We excluded all patients who received Paxlovid before t0, and those who were hospitalized between t0 and time of treatment assignment (*t_A_*) (prescription of Paxlovid). This period, a patient’s “immortal time”, may bias estimates in favor of a lower risk in the treatment group, even in the absence of a true effect. ^26^ To quantify the bias due to the presence of immortal time, in this sensitivity analysis we included treated patients who were hospitalized on or between *t*_0_ and *t_A_*. This represents a “worst-case scenario”, and provides a conservative estimate of the treatment effect taking into account the potential immortal time bias.

## Declarations

### Ethics approval and Consent to Participate

The N3C data transfer to NCATS is performed under a Johns Hopkins University Reliance Protocol # IRB00249128 or individual site agreements with NIH. The N3C Data Enclave is managed under the authority of the NIH; information can be found at https://ncats.nih.gov/n3c/resources. The work was performed under DUR RP-5677B5.

### Data Availability

All data is available in the N3C Data Enclave to those with an approved protocol and data use request from an institutional review board. Data access is governed under the authority of the National Institutes of Health; more information on accessing the data can be found at https://covid.cd2h.org/for-researchers. See Haendel et. al. for additional detail on how data is ingested, managed, and protected within the N3C Data Enclave. ^34^

### Code Availability

The N3C Data Enclave is available for public use. To access data used within this manuscript, institutions must have a signed Data Use Agreement executed with the U.S. National Center for Advancing Translational Sciences (NCATS) and their investigators must complete mandatory training and must submit a Data Use Request (DUR) to N3C. To request N3C data access, researchers must follow instructions at https://covid.cd2h.org/onboarding. Code is available to those with valid login credentials for the N3C Data Enclave. It was written for use in the enclave on the Palantir Foundry platform, where the analysis can be reproduced by researchers. It can be exported for review upon request. ^46^

## Acknowledgements

Authorship was determined using ICMJE recommendations. The content is solely the responsibility of the authors and does not necessarily represent the official views of the National Institutes of Health or N3C.

## N3C Attribution

The analyses described in this publication were conducted with data or tools accessed through the NCATS N3C Data Enclave (covid.cd2h.org/enclave) and supported by CD2H - The National COVID Cohort Collaborative (N3C) IDeA CTR Collaboration 3U24TR002306-04S2 NCATS U24 TR002306. This research was possible because of the patients whose information is included within the data from participating organizations (covid.cd2h.org/dtas) and the organizations and scientists (covid.cd2h.org/duas) who have contributed to the on-going development of this community resource. ^34^

### We gratefully acknowledge the following core contributors to N3C

Adam B. Wilcox, Adam M. Lee, Alexis Graves, Alfred (Jerrod) Anzalone, Amin Manna, Amit Saha, Amy Olex, Andrea Zhou, Andrew E. Williams, Andrew Southerland, Andrew T. Girvin, Anita Walden, Anjali A. Sharathkumar, Benjamin Amor, Benjamin Bates, Brian Hendricks, Brijesh Patel, Caleb Alexander, Carolyn Bramante, Cavin Ward-Caviness, Charisse Madlock-Brown, Christine Suver, Christopher Chute, Christopher Dillon, Chunlei Wu, Clare Schmitt, Cliff Takemoto, Dan Housman, Davera Gabriel, David A. Eichmann, Diego Mazzotti, Don Brown, Eilis Boudreau, Elaine Hill, Elizabeth Zampino, Emily Carlson Marti, Emily R. Pfaff, Evan French, Farrukh M Koraishy, Federico Mariona, Fred Prior, George Sokos, Greg Martin, Harold Lehmann, Heidi Spratt, Hemalkumar Mehta, Hongfang Liu, Hythem Sidky, J.W. Awori Hayanga, Jami Pincavitch, Jaylyn Clark, Jeremy Richard Harper, Jessica Islam, Jin Ge, Joel Gagnier, Joel H. Saltz, Joel Saltz, Johanna Loomba, John Buse, Jomol Mathew, Joni L. Rutter, Julie A. McMurry, Justin Guinney, Justin Starren, Karen Crowley, Katie Rebecca Bradwell, Kellie M. Walters, Ken Wilkins, Kenneth R. Gersing, Kenrick Dwain Cato, Kimberly Murray, Kristin Kostka, Lavance Northington, Lee Allan Pyles, Leonie Misquitta, Lesley Cottrell, Lili Portilla, Mariam Deacy, Mark M. Bissell, Marshall Clark, Mary Emmett, Mary Morrison Saltz, Matvey B. Palchuk, Melissa A. Haendel, Meredith Adams, Meredith Temple-O’Connor, Michael G. Kurilla, Michele Morris, Nabeel Qureshi, Nasia Safdar, Nicole Garbarini, Noha Sharafeldin, Ofer Sadan, Patricia A. Francis, Penny Wung Burgoon, Peter Robinson, Philip R.O. Payne, Rafael Fuentes, Randeep Jawa, Rebecca Erwin-Cohen, Rena Patel, Richard A. Moffitt, Richard L. Zhu, Rishi Kamaleswaran, Robert Hurley, Robert T. Miller, Saiju Pyarajan, Sam G. Michael, Samuel Bozzette, Sandeep Mallipattu, Satyanarayana Vedula, Scott Chapman, Shawn T. O’Neil, Soko Setoguchi, Stephanie S. Hong, Steve Johnson, Tellen D. Bennett, Tiffany Callahan, Umit Topaloglu, Usman Sheikh, Valery Gordon, Vignesh Subbian, Warren A. Kibbe, Wenndy Hernandez, Will Beasley, Will Cooper, William Hillegass, Xiaohan Tanner Zhang. Details of contributions available at covid.cd2h.org/core-contributors

## Data Partners

The following institutions whose data is released or pending:

### Available

Advocate Health Care Network — UL1TR002389: The Institute for Translational Medicine (ITM) • Boston University Medical Campus — UL1TR001430: Boston University Clinical and Translational Science Institute • Brown University — U54GM115677: Advance Clinical Translational Research (Advance-CTR) • Carilion Clinic — UL1TR003015: iTHRIV Integrated Translational health Research Institute of Virginia • Charleston Area Medical Center U54GM104942: West Virginia Clinical and Translational Science Institute (WVCTSI) • Children’s Hospital Colorado — UL1TR002535: Colorado Clinical and Translational Sciences Institute Columbia University Irving Medical Center — UL1TR001873: Irving Institute for Clinical and Translational Research • Duke University — UL1TR002553: Duke Clinical and Translational Science Institute • George Washington Children’s Research Institute — UL1TR001876: Clinical and Translational Science Institute at Children’s National (CTSA-CN) • George Washington University — UL1TR001876: Clinical and Translational Science Institute at Children’s National (CTSA-CN) • Indiana University School of Medicine — UL1TR002529: Indiana Clinical and Translational Science Institute • Johns Hopkins University — UL1TR003098: Johns Hopkins Institute for Clinical and Translational Research • Loyola Medicine — Loyola University Medical Center • Loyola University Medical Center — UL1TR002389: The Institute for Translational Medicine (ITM) • Maine Medical Center — U54GM115516: Northern New England Clinical & Translational Research (NNE-CTR) Network • Massachusetts General Brigham — UL1TR002541: Harvard Catalyst • Mayo Clinic Rochester — UL1TR002377: Mayo Clinic Center for Clinical and Translational Science (CCaTS) • Medical University of South Carolina — UL1TR001450: South Carolina Clinical & Translational Research Institute (SCTR) • Montefiore Medical Center UL1TR002556: Institute for Clinical and Translational Research at Einstein and Montefiore • Nemours — U54GM104941: Delaware CTR ACCEL Program • NorthShore University HealthSystem — UL1TR002389: The Institute for Translational Medicine (ITM) • Northwestern University at Chicago — UL1TR001422: Northwestern University Clinical and Translational Science Institute (NUCATS) • OCHIN — INV-018455: Bill and Melinda Gates Foundation grant to Sage Bionetworks • Oregon Health & Science University — UL1TR002369: Oregon Clinical and Translational Research Institute • Penn State Health Milton S. Hershey Medical Center — UL1TR002014: Penn State Clinical and Translational Science Institute • Rush University Medical Center — UL1TR002389: The Institute for Translational Medicine (ITM) • Rutgers, The State University of New Jersey — UL1TR003017: New Jersey Alliance for Clinical and Translational Science • Stony Brook University — U24TR002306 • The Ohio State University — UL1TR002733: Center for Clinical and Translational Science • The State University of New York at Buffalo — UL1TR001412: Clinical and Translational Science Institute • The University of Chicago — UL1TR002389: The Institute for Translational Medicine (ITM) • The University of Iowa — UL1TR002537: Institute for Clinical and Translational Science • The University of Miami Leonard M. Miller School of Medicine — UL1TR002736: University of Miami Clinical and Translational Science Institute • The University of Michigan at Ann Arbor — UL1TR002240: Michigan Institute for Clinical and Health Research • The University of Texas Health Science Center at Houston — UL1TR003167: Center for Clinical and Translational Sciences (CCTS) • The University of Texas Medical Branch at Galveston — UL1TR001439: The Institute for Translational Sciences • The University of Utah — UL1TR002538: Uhealth Center for Clinical and Translational Science • Tufts Medical Center — UL1TR002544: Tufts Clinical and Translational Science Institute • Tulane University UL1TR003096: Center for Clinical and Translational Science • University Medical Center New Orleans — U54GM104940: Louisiana Clinical and Translational Science (LA CaTS) Center • University of Alabama at Birmingham — UL1TR003096: Center for Clinical and Translational Science • University of Arkansas for Medical Sciences — UL1TR003107: UAMS Translational Research Institute • University of Cincinnati — UL1TR001425: Center for Clinical and Translational Science and Training • University of Colorado Denver, Anschutz Medical Campus — UL1TR002535: Colorado Clinical and Translational Sciences Institute • University of Illinois at Chicago — UL1TR002003: UIC Center for Clinical and Translational Science • University of Kansas Medical Center — UL1TR002366: Frontiers: University of Kansas Clinical and Translational Science Institute • University of Kentucky — UL1TR001998: UK Center for Clinical and Translational Science • University of Massachusetts Medical School Worcester — UL1TR001453: The UMass Center for Clinical and Translational Science (UMCCTS) • University of Minnesota UL1TR002494: Clinical and Translational Science Institute • University of Mississippi Medical Center — U54GM115428: Mississippi Center for Clinical and Translational Research (CCTR) • University of Nebraska Medical Center — U54GM115458: Great Plains IDeA-Clinical & Translational Research • University of North Carolina at Chapel Hill — UL1TR002489: North Carolina Translational and Clinical Science Institute • University of Oklahoma Health Sciences Center U54GM104938: Oklahoma Clinical and Translational Science Institute (OCTSI) • University of Rochester — UL1TR002001: UR Clinical & Translational Science Institute • University of Southern California — UL1TR001855: The Southern California Clinical and Translational Science Institute (SC CTSI) • University of Vermont — U54GM115516: Northern New England Clinical & Translational Research (NNE-CTR) Network • University of Virginia — UL1TR003015: iTHRIV Integrated Translational health Research Institute of Virginia • University of Washington UL1TR002319: Institute of Translational Health Sciences • University of Wisconsin-Madison UL1TR002373: UW Institute for Clinical and Translational Research • Vanderbilt University Medical Center — UL1TR002243: Vanderbilt Institute for Clinical and Translational Research • Virginia Commonwealth University — UL1TR002649: C. Kenneth and Dianne Wright Center for Clinical and Translational Research • Wake Forest University Health Sciences — UL1TR001420: Wake Forest Clinical and Translational Science Institute • Washington University in St. Louis — UL1TR002345: Institute of Clinical and Translational Sciences • Weill Medical College of Cornell University — UL1TR002384: Weill Cornell Medicine Clinical and Translational Science Center West Virginia University — U54GM104942: West Virginia Clinical and Translational Science Institute (WVCTSI)

### Submitted

Icahn School of Medicine at Mount Sinai — UL1TR001433: ConduITS Institute for Translational Sciences • The University of Texas Health Science Center at Tyler — UL1TR003167: Center for Clinical and Translational Sciences (CCTS) • University of California, Davis — UL1TR001860: UCDavis Health Clinical and Translational Science Center • University of California, Irvine — UL1TR001414: The UC Irvine Institute for Clinical and Translational Science (ICTS) • University of California, Los Angeles — UL1TR001881: UCLA Clinical Translational Science Institute • University of California, San Diego — UL1TR001442: Altman Clinical and Translational Research Institute • University of California, San Francisco — UL1TR001872: UCSF Clinical and Translational Science Institute

### Pending

Arkansas Children’s Hospital — UL1TR003107: UAMS Translational Research Institute • Baylor College of Medicine — None (Voluntary) • Children’s Hospital of Philadelphia — UL1TR001878: Institute for Translational Medicine and Therapeutics • Cincinnati Children’s Hospital Medical Center — UL1TR001425: Center for Clinical and Translational Science and Training • Emory University — UL1TR002378: Georgia Clinical and Translational Science Alliance • HonorHealth — None (Voluntary) • Loyola University Chicago — UL1TR002389: The Institute for Translational Medicine (ITM) • Medical College of Wisconsin — UL1TR001436: Clinical and Translational Science Institute of Southeast Wisconsin • MedStar Health Research Institute — UL1TR001409: The Georgetown-Howard Universities Center for Clinical and Translational Science (GHUCCTS) • MetroHealth — None (Voluntary) • Montana State University U54GM115371: American Indian/Alaska Native CTR • NYU Langone Medical Center — UL1TR001445: Langone Health’s Clinical and Translational Science Institute • Ochsner Medical Center — U54GM104940: Louisiana Clinical and Translational Science (LA CaTS) Center • Regenstrief Institute — UL1TR002529: Indiana Clinical and Translational Science Institute • Sanford Research — None (Voluntary) • Stanford University — UL1TR003142: Spectrum: The Stanford Center for Clinical and Translational Research and Education • The Rockefeller University — UL1TR001866: Center for Clinical and Translational Science • The Scripps Research Institute — UL1TR002550: Scripps Research Translational Institute • University of Florida UL1TR001427: UF Clinical and Translational Science Institute • University of New Mexico Health Sciences Center — UL1TR001449: University of New Mexico Clinical and Translational Science Center • University of Texas Health Science Center at San Antonio — UL1TR002645: Institute for Integration of Medicine and Science • Yale New Haven Hospital — UL1TR001863: Yale Center for Clinical Investigation

## Author Contributions

A.B., A.J.P., X.X., M.D.B., C.M.B. performed the data analysis. K.J.W. advised in the design of the statistical analysis. H.D. and M.F. provided substantive input to the interpretation and conclusions as patient representatives, and G.C.A. provided substantive input as a clinician. A.B., A.J.P., X.X., M.D.B., G.C.A, R.F.C., E.H., E.P.K., H.B.M., K.J.W., R.M., C.G.C., and E.R.P. wrote and revised the manuscript. A.B., A.J.P., M.D.B., C.M.B., R.M. and E.R.P. contributed significantly to cohort definition and creation. M.H. and C.G.C. supervised the study.

## Supplementary Tables and Figures

**Fig. 1.**
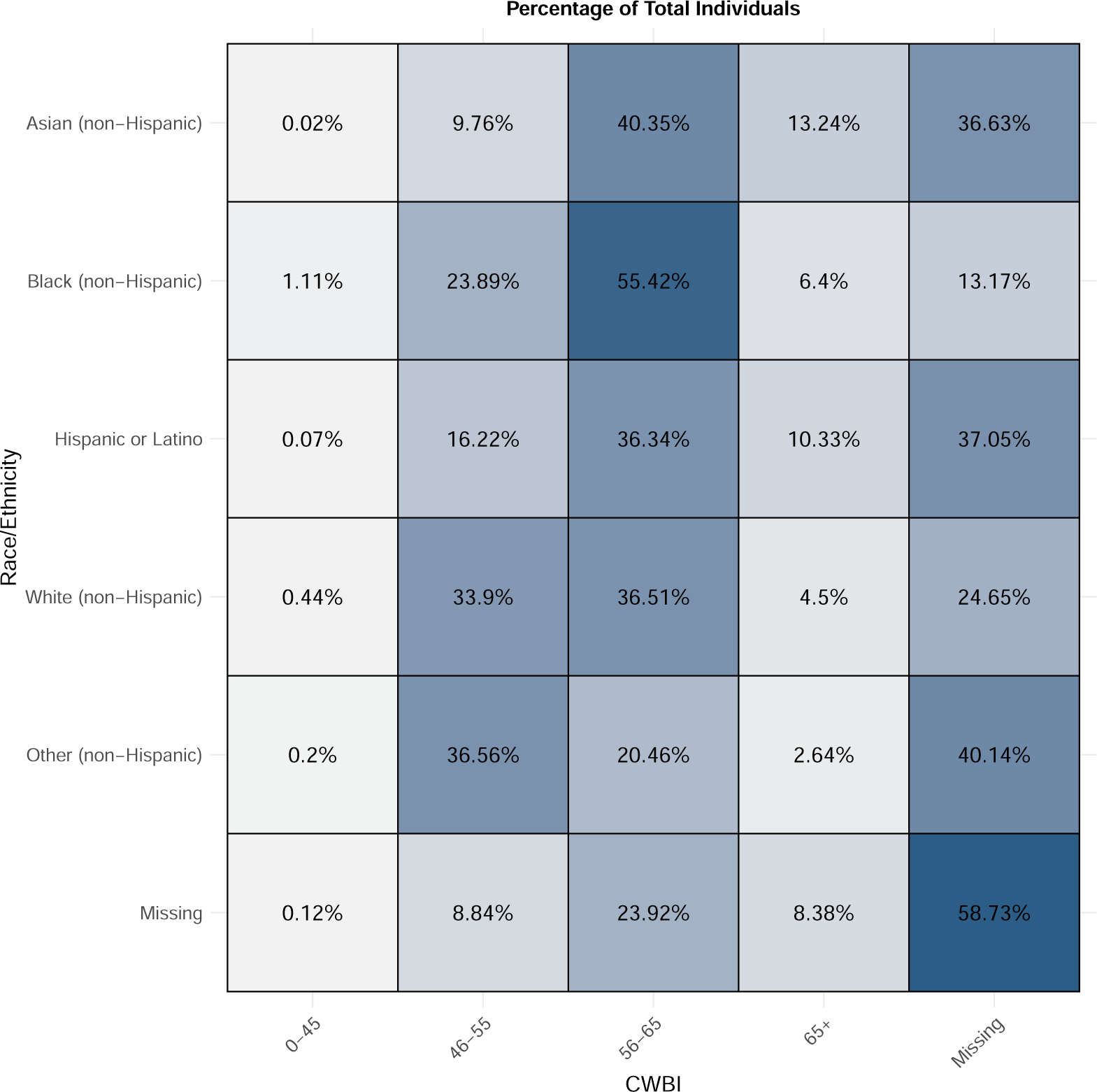
Study Cohort and Flow of Emulated Trial **2,086,107** patients with a positive PCR or antigen test for SARS-CoV-2 between 22 December 2021 and 31 December 2022

**Table 1.** Sensitivity analysis examining the effects of immortal time bias, and missingness in the ZIP Code-level Community Wellbeing Index (CWBI)

|  | *Dependent variable: Hospitalization* |  |  |
| --- | --- | --- | --- |
|  | Not accounting for<br>Immortal Time Bias | Dropping CWBI<br>covariate | Dropping sites with<br>missing CWBI |
|  | OR (95% CI) | OR (95% CI) | OR (95% CI) |
| Paxlovid Treatment |  |  |  |
| No (control) | *ref* | *ref* | *ref* |
| Yes (treatment) | 0.38 (0.31-0.46)*** | 0.37 (0.31-0.45)*** | 0.37 (0.29-0.47)*** |
| *Observations* | 410,716 | 455,504 | 272,133 |
| *Log Likelihood* | -29151.5 | -29618.4 | -20744.2 |
| *Akaike Inf. Crit.* | 58376.9 | 59296.7 | 41562.4 |

Note: $p < .05^*$ , $p < .01^{**}$ , $p < .001^{***}$

## References

[1] WHO coronavirus (COVID-19) dashboard. https://covid19.who.int/. Accessed: 2023-3-26.

[2] Center for Drug Evaluation & Research. FDA updates on paxlovid for health care providers. https://www.fda.gov/drugs/news-events-human-drugs/fda-updates-paxlovid-health-care-providers. Accessed: 2023-3-26.

[3] Hammond, J. et al. Oral nirmatrelvir for High-Risk, nonhospitalized adults with covid-19. N. Engl. J. Med. 386, 1397–1408 (2022).

[4] Malden, D. E. et al. Hospitalization and Emergency Department Encounters for COVID-19 After Paxlovid Treatment - California, December 2021-May 2022. MMWR. Morbidity and mortality weekly report 71, 830–833 (2022).

[5] Najjar-Debbiny, R. et al. Effectiveness of paxlovid in reducing severe coronavirus disease 2019 and mortality in High-Risk patients. Clin. Infect. Dis. 76, e342–e349 (2023).

[6] Yip, T. C.-F. et al. Impact of the use of oral antiviral agents on the risk of hospitalization in community coronavirus disease 2019 patients (COVID-19). Clin. Infect. Dis. 76, e26–e33 (2023).

[7] Klein, E. J., Hardesty, A., Vieira, K. & Farmakiotis, D. Use of anti-spike monoclonal antibodies in kidney transplant recipients with COVID-19: Efficacy, ethnic and racial disparities. Am. J. Transplant 22, 640–645 (2022).

[8] Wiltz, J. L. et al. Racial and ethnic disparities in receipt of medications for treatment of COVID-19 - united states, march 2020-august 2021. MMWR Morb. Mortal. Wkly. Rep. 71, 96–102 (2022).

[9] Wu, E.-L. et al. Disparities in COVID-19 monoclonal antibody delivery: a retrospective cohort study. J. Gen. Intern. Med. 37, 2505–2513 (2022).

[10] Boehmer, T. K. et al. Racial and ethnic disparities in outpatient treatment of COVID-19 - united states, January-July 2022. MMWR Morb. Mortal. Wkly. *Rep.* 71, 1359–1365 (2022).

[11] Herńan, M. A. & Robins, J. M. Using big data to emulate a target trial when a randomized trial is not available. Am. J. Epidemiol. 183, 758–764 (2016).

[12] Chin, T. et al. US-county level variation in intersecting individual, household and community characteristics relevant to COVID-19 and planning an equitable response: a cross-sectional analysis. BMJ Open 10, e039886 (2020).

[13] Garg, S. et al. Hospitalization rates and characteristics of patients hospitalized with Laboratory-Confirmed coronavirus disease 2019 - COVID-NET, 14 states, march 1-30, 2020. MMWR Morb. Mortal. Wkly*. Rep.* 69, 458–464 (2020).

[14] van Dorn, A., Cooney, R. E. & Sabin, M. L. COVID-19 exacerbating inequalities in the US. Lancet 395, 1243–1244 (2020).

[15] Raifman, M. A. & Raifman, J. R. Disparities in the population at risk of severe illness from COVID-19 by Race/Ethnicity and income. Am. J. Prev. Med. 59, 137–139 (2020).

[16] Dyer, O. Covid-19: Black people and other minorities are hardest hit in US. BMJ 369, m1483 (2020).

[17] Webb Hooper, M., Ńapoles, A. M. & Pérez-Stable, E. J. COVID-19 and Racial/Ethnic disparities. JAMA 323, 2466–2467 (2020).

[18] Dryden-Peterson, S. et al. Nirmatrelvir plus ritonavir for early COVID-19 in a large U.S. health system : A Population-Based cohort study. Ann. Intern. Med. 176, 77–84 (2023).

[19] Gupta, S. et al. Association between early treatment with tocilizumab and mortality among critically ill patients with COVID-19. JAMA Intern. Med. 181, 41–51 (2021).

[20] Petito, L. C. et al. Estimates of overall survival in patients with cancer receiving different treatment regimens: Emulating hypothetical target trials in the surveillance, epidemiology, and end results (SEER)-Medicare linked database. JAMA Netw Open 3, e200452 (2020).

[21] Dickerman, B. A., Garćıa-Albéniz, X., Logan, R. W., Denaxas, S. & Herńan, M. A. Avoidable flaws in observational analyses: an application to statins and cancer. Nat. Med. 25, 1601–1606 (2019).

[22] Admon, A. J. et al. Emulating a novel clinical trial using existing observational data. predicting results of the PreVent study. Ann. Am. Thorac. Soc. 16, 998–1007 (2019).

[23] van Walraven, C., Bennett, C. & Forster, A. J. Administrative database research infrequently used validated diagnostic or procedural codes. J. Clin. Epidemiol. 64, 1054–1059 (2011).

[24] Stuart, E. A. Matching methods for causal inference: A review and a look forward. Stat. Sci. 25, 1–21 (2010).

[25] Imai, K., King, G. & Stuart, E. A. Misunderstandings between experimentalists and observationalists about causal inference. J. R. Stat. Soc. Ser. A Stat. Soc. 171, 481–502 (2008).

[26] Herńan, M. A., Sauer, B. C., Herńandez-Díaz, S., Platt, R. & Shrier, I. Specifying a target trial prevents immortal time bias and other self-inflicted injuries in observational analyses. J. Clin. Epidemiol. 79, 70–75 (2016).

[27] Caniglia, E. C. et al. Emulating target trials to avoid immortal time biasan application to antibiotic initiation and preterm delivery. Epidemiology (2023).

[28] Wells, B. J., Chagin, K. M., Nowacki, A. S. & Kattan, M. W. Strategies for handling missing data in electronic health record derived data. EGEMS (Wash DC) 1, 1035 (2013).

[29] Lin, K. J. et al. Out-of-system care and recording of patient characteristics critical for comparative effectiveness research. Epidemiology 29, 356–363 (2018).

[30] Wang, L. E., Shaw, P. A., Mathelier, H. M., Kimmel, S. E. & French, B. Evaluating Risk-Prediction models using data from electronic health records. Ann. Appl. Stat. 10, 286–304 (2016).

[31] CDC. Interim clinical considerations for COVID-19 treatment in outpatients. https://www.cdc.gov/coronavirus/2019-ncov/hcp/clinical-care/outpatient-treatment-overview.html(2023). Accessed: 2023-3-26.

[32] SPH and sharecare release community Well-Being rankings. https://www.bu.edu/sph/news/articles/2020/sph-and-sharecare-release-community-well-being-rankings/. Accessed: 2023-3-30.

[33] Herńan, M. A., Wang, W. & Leaf, D. E. Target trial emulation: A framework for causal inference from observational data. JAMA 328, 2446–2447 (2022).

[34] Haendel, M. A. et al. The national COVID cohort collaborative (N3C): Rationale, design, infrastructure, and deployment. J. Am. Med. Inform. Assoc. 28, 427–443 (2021).

[35] Pfaff, E. R. et al. Synergies between centralized and federated approaches to data quality: a report from the national COVID cohort collaborative. J. Am. Med. Inform. Assoc. 29, 609–618 (2022).

[36] Phenotype Data Acquisition: The repository for code and documentation produced by the N3C phenotype and data acquisition workstream.

[37] British Medical Journal Publishing Group. Strengthening the reporting of observational studies in epidemiology (STROBE) statement: guidelines for reporting observational studies. BMJ 335 (2007).

[38] Howard-Jones, A. R., et al. COVID-19 in children. II: Pathogenesis, disease spectrum and management. J. Paediatr. Child Health 58, 46–53 (2022).

[39] Bose-Brill, S., et al. Pediatric Nirmatrelvir/Ritonavir prescribing patterns during the COVID-19 pandemic. medRxiv (2022).

[40] Hripcsak, G. et al. Observational health data sciences and informatics (OHDSI): Opportunities for observational researchers. Stud. Health Technol. Inform. 216, 574–578 (2015).

[41] Iacus, S. M., King, G. & Porro, G. Causal inference without balance checking: Coarsened exact matching. Polit. Anal. 20, 1–24 (2012).

[42] Gory, J. J., Craigmile, P. F. & MacEachern, S. N. A class of generalized linear mixed models adjusted for marginal interpretability. Stat. Med. 40, 427–440 (2021).

[43] Evaluation of protease inhibition for COVID-19 in Standard-Risk patients (EPIC-SR). https://clinicaltrials.gov/ct2/show/NCT05011513. Accessed: 2023-3-31.

[44] CDC. Data definitions for COVID-19 vaccinations in the united states. https://www.cdc.gov/coronavirus/2019-ncov/vaccines/reporting-vaccinations.html (2023). Accessed: 2023-2-20.

[45] Brannock, M. D., et al. Long COVID risk and Pre-COVID vaccination: An EHR-Based cohort study from the RECOVER program. medRxiv (2022).

[46] Palantir foundry. https://www.palantir.com/platforms/foundry/. Accessed: 2023-1-9.

